# Proprioceptive Acuity of the Ankle is Higher in Plantarflexion than in Dorsiflexion

**DOI:** 10.1101/2025.03.24.25324535

**Authors:** Yung-Tze Lee, Farina Mirbagheri, Xinyi Zhou, Jürgen Konczak

**Affiliations:** Human Sensorimotor Control Laboratory, School of Kinesiology, University of Minnesota, Minneapolis, MN, USA

**Author notes:** Corresponding author: Jürgen Konczak, PhD. University of Minnesota, School of Kinesiology. These authors contributed equally (Yung-Tze Lee, Farina Mirbagheri).

**Keywords:** human, position sense, proprioceptive acuity, proprioceptive discrimination

## Abstract

Intact ankle proprioception is essential for the control of balance and gait. This study determined ankle position sense acuity for plantarflexion and dorsiflexion. In two separate assessments, the right ankle of 30 healthy young adults was passively rotated from neutral joint position to a 15° reference position and a smaller comparison position in either plantar- or dorsiflexion. Subsequently, participants verbally indicated which position felt more flexed. After 25 trials, a psychometric function was fitted to the respective response-stimulus size difference data for each participant and two outcome measures were derived: a Just-Noticeable-Difference (JND) threshold as a measure of systematic error and an Uncertainty Area (UA) indicating random error. Analysis showed that mean JND threshold and median UA were both significantly higher in dorsiflexion when compared to plantarflexion (p = 0.008, d = 0.52; p = 0.001, *r*_*b*_= 0.58). These findings indicate that ankle proprioceptive acuity is not uniform for sagittal plane ankle motion but is higher for plantarflexion. We discuss differences in plantar and dorsiflexor muscle mechanoreceptor density and central proprioceptive signal processing as possible reasons for the observed differences in acuity and highlight the importance of understanding movement-specific proprioceptive acuity for designing effective rehabilitation protocols.

## Introduction

Proprioception refers to the sense of body position, motion and force (Goldscheider, 1898; Sherrington, 1907). Proprioceptive mechanoreceptors embedded in muscles, ligaments, and tendons relay information regarding muscle tension, tissue strain, and stretch velocity, which are critical for volitional and reflexive motor control. With respect to balance and gait, ankle proprioception is crucial because proprioceptive afferents provide information about ankle position and motion that are used by the nervous system for the control of balance and to produce fast postural responses to mechanical perturbations (Allum et al., 1998; Goble et al., 2011; Gordon et al., 1995; Hubbuch et al., 2015). Conversely, peripheral or central nervous system diseases such as diabetes-induced lower limb neuropathy or Parkinson’s disease are associated with impairments in gait and loss of balance (Henry & Baudry, 2019; Horak, 2006).

The accuracy of the proprioceptive sense is typically related to motor function and may be similar between homologous joints (e.g., left and right ankle) (Sertic et al., 2024), may differ between non-homologous joints, or between the various degrees-of-freedom within a given joint (e.g., wrist flexion/extension vs. ad/abduction) (Cappello et al., 2015). Anatomical and biomechanical evidence suggests that proprioceptive acuity might also differ between plantar- and dorsiflexion due to variations in muscle length, tendon stretch, and joint mechanics (Brockett & Chapman, 2016; Lai et al., 2016). However, at present, no empirical evidence exists indicating differences between or within the degrees-of-freedom (DoF) of the ankle joint.

To bridge this knowledge gap, this study systematically investigated differences in ankle position sense between dorsiflexion and plantarflexion in healthy younger adults using the Ankle Proprioceptive Acuity System (APAS). Specifically, we focused on passive ankle position sense, which refers to the individual’s ability to differentiate between two ankle positions (Goldscheider, 1898; Winter et al., 2022) while a joint is passively rotated. It relies on the somatosensory cortical processing of proprioceptive afferents. In contrast, active position sense requires muscle activation of the perceiver and involves sensory as well as sensorimotor motor integration processes (Elangovan et al., 2014). We employed a sensory psychophysics approach examining the relation between a specific ankle joint position and the evoked perception (Elangovan et al., 2014; Gescheider, 1985). The method yields two measures reflecting proprioceptive acuity, the discrimination or just-noticeable-difference threshold, which represents a measure of perceptual bias or systematic error, and the uncertainty area reflecting the consistency of perception over repeated exposure to the same joint position. It constitutes a measure of precision or variable error.

## Methods

### Participants

Thirty young adults (15 females; age range: 18 – 30 years, mean: 25.1 ± 2.3 years) participated in the study. Based on previous data of healthy young adults using the same device (Sertic et al., 2024), an a-priori power analysis was performed (power = 0.8, α = 0.05, d = 0.55) that yielded n = 29 as necessary sample size.

The inclusion criterion for recruitment was an age range between 18 to 30 years. The exclusion criteria were: 1) prior history of a neurological disorder affecting movement or somatosensation, 2) history of lower fractures, 3) present lower body injuries, and 4) amputation or joint replacement of any part of the leg. Prior to consenting, potential participants answered a screening questionnaire to ensure eligibility based on the inclusion/exclusion criteria. In terms of ethnicity, participants identified as Asian (n = 19), White (n = 7), Hispanic (n = 3), or African American (n = 1). Written informed consent was secured from each participant before the study began. The study protocol was approved by the University of Minnesota Institutional Review Board (STUDY00021532).

### Testing Apparatus

The Ankle Proprioceptive Acuity System (APAS) was used to measure ankle proprioception (Mahnan et al., 2020) (**Figure 1A**). This apparatus allows for the passive rotation of the ankle joint around its mediolateral axis (plantar/dorsiflexion). Participants positioned their right foot on the platform, aligning the lateral malleolus of the tibia with the platform’s rotational axis. The foot was secured to the platform with a Velcro strap to ensure a stable foot position. Moving the handle of the APAS system, the experimenter manually rotated the foot slowly (∼ 6°/s) from an initial to a target position. To ensure precise and repeatable displacement of the joint, metal pins were inserted into designated holes on a pegboard as mechanical stops. The holes were spaced in such a way that the minimal difference between two distinct ankle positions was 0.1°. The validity and efficacy of the APAS system in measuring ankle position sense acuity was established previously (Mahnan et al., 2020).

**Figure 1.**
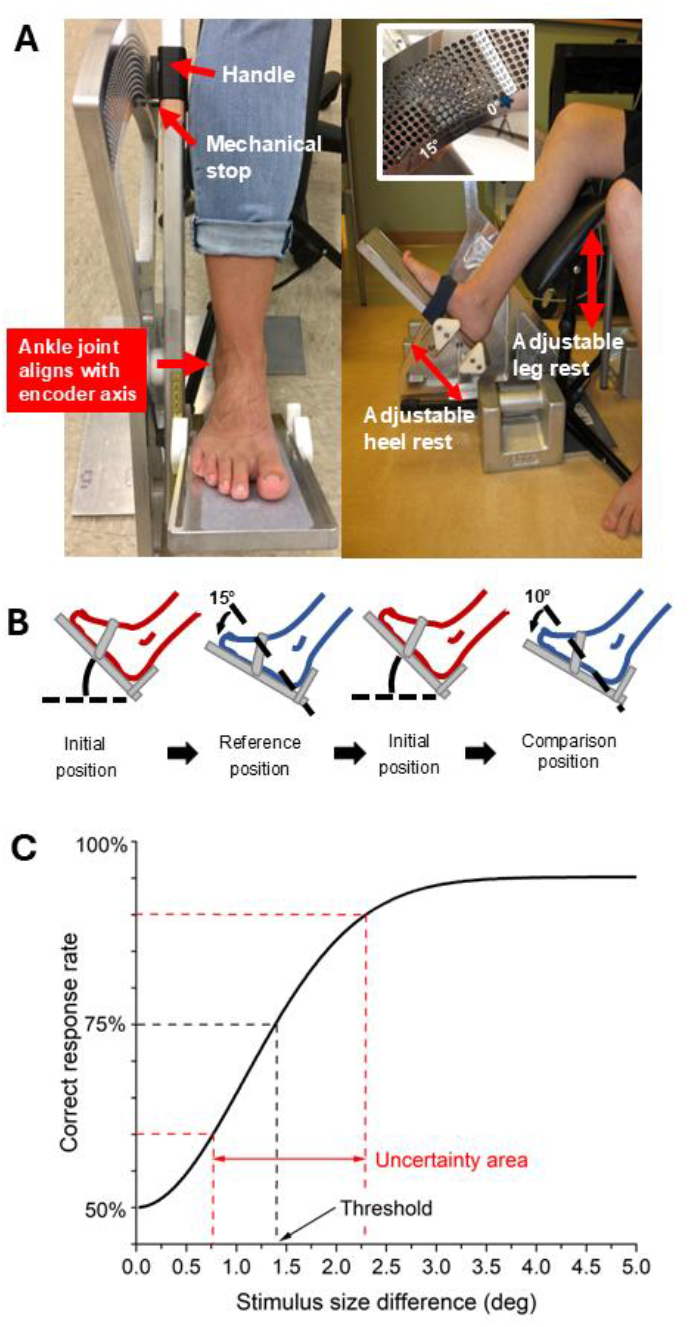
**A)** Image of the Ankle Proprioceptive Acuity System. Leg rest and heel rest are set to align the ankle with the central axis of foot platform. Metal pins inserted in the pegboard holes serve as mechanical stops to assure the accuracy of specific start and target position. **B)** Process diagram of a single trial for the protocol of plantarflexion. The foot is in an initial neutral position. The foot is rotated to the *reference* position, which is held for 2 seconds and then returns to the initial position. Subsequently, the experimenter rotates the foot to the *comparison* position and then back to the initial position. Finally, the participant verbally indicates which position was perceived as being closer to the ground. **C)** Example of a psychometric function derived from the response – stimulus size difference data. The JND threshold corresponds to the stimulus size difference at which perceiver correctly discriminated between reference and comparison position of the ankle with 75% accuracy. The Uncertainty Area corresponds to the range of stimulus size differences at 60% and 90% correct response rate. Figure 1A is modified from Mahnan et al. (2020) with permission.

### Procedure

During the testing, participants sat blindfolded, with their feet positioned on the APAS platform. The shank rested on a leg support to assure an unweighted position so that differences in loading the sole of the foot could not be used as a sensory cue to judge ankle position (**Figure 1A**).

During testing, electromyographic activity of tibialis anterior, soleus, and gastrocnemius were monitored online by the experimenters to assure that the participant did not actively move the foot. If such activation occurred, the trial was repeated. Participants were allowed to take intermittent breaks after every 7 trials, with an additional break between each test protocol. Two test protocols were applied, one for examining plantarflexed and one for dorsiflexed positions. Each protocol comprised 25 trials. The order of protocols (1^st^ or 2^nd^) was randomized between subjects to control for possible order effects.

We employed a psychophysical two-alternative forced-choice sensory paradigm (Gescheider, 1985). For the assessment of plantarflexion, each trial consisted of the experimenter rotating the participant’s ankle from an initial position (5° dorsiflexion from neutral = 90°) to two distinct plantarflexed positions: a *reference* position (15°) and a *comparison* position (< 15°) (**Figure 1B**). The foot was maintained at each position for 2 seconds. After experiencing the second position, participants verbally indicated whether their toes were closer to the ground in the ‘first’ or ‘second’ position. After each trial, the comparison position of the subsequent trial was determined using a Bayesian psi-marginal algorithm based on the prior angular difference between reference and comparison position and the respective participant response (Prins, 2013). This procedure assured fast convergence of the trials towards the perceptual discrimination threshold. For each trial, the order of reference and comparison position was randomized to control for order effects. Participants did not receive feedback about the correctness of their responses to eliminate any learning effects over successive trials.

For assessing dorsiflexion, the evaluation process was identical, except that the initial ankle joint position was set to 10° plantarflexion from neutral and the *reference* was a 15° dorsiflexed position, and the *comparison* position was < 15°.

### Measurements

After the completion of testing, a logistic Weibull function was fitted to the obtained verbal response – position difference (reference – comparison) data for each test of each participant. Based on this psychometric function, two outcome measures of position sense acuity were determined: 1) the *just-noticeable-difference (JND) threshold*, which refers to the perceived difference in ankle position at the 75% correct response rate, and 2) the *Uncertainty Area* (UA), which is the angular position difference between the 60-90% correct response rate. The JND threshold represents systematic error and is a measure of perceptual *bias*. The UA represents the random error across repeated perceptions and is a measure of perceptual *precision*. Both variables reflect a person’s perceptual accuracy in discriminating between ankle joint positions. A lower JND threshold and a lower UA value indicate a higher acuity in position sensing (**Figure 1C**).

### Data Analysis

A Shapiro-Wilk test was performed to evaluate the normality assumption for JND threshold and UA data sets. The results showed that the JND threshold data were normally distributed. The UA data were not normally distributed. Given that the JND threshold was normally distributed, a paired t-test was used to compare the difference in plantarflexion and dorsiflexion. For non-normally distributed UA data, we applied a Wilcoxon signed-rank test. UA values were classified as outliers if they fell below the 5th or above the 95th percentile. After conducting statistical analyses with and without these outliers and observing no difference in the results, we decided to retain the outliers for UA values. Effect size was determined using Cohen’s d for the normally distributed JND threshold data and the rank-biserial correlation (*r*_*b*_) is reported for the non-normally distributed UA data (Cohen, 1988; Fritz et al., 2012). The significance level for all tests was set at p < 0.05. All statistical analyses were conducted using R software version 4.2.2.

## Results

### Differences in JND Threshold between Plantarflexion and Dorsiflexion

To assess systematic differences in proprioceptive bias for plantar-versus dorsiflexed ankle position, we analyzed the individual JND threshold data of all participants. As shown in **Figure 2A**, a total of 21/30 (70%) participants exhibited a higher threshold in dorsiflexion when compared to plantarflexion. A corresponding paired t-test indicated that the mean ankle JND threshold for plantarflexion was significantly lower when compared to dorsiflexion (*Mean* plantarflexion: 1.3°, SD: 0.7°; *Mean* dorsiflexion: 1.8°, SD: 0.7°; *t*_(29)_ = 2.85, *p* = 0.008) with a moderate to strong effect size (*d* = 0.52) (**Figure 2B)**.

**Figure 2.**
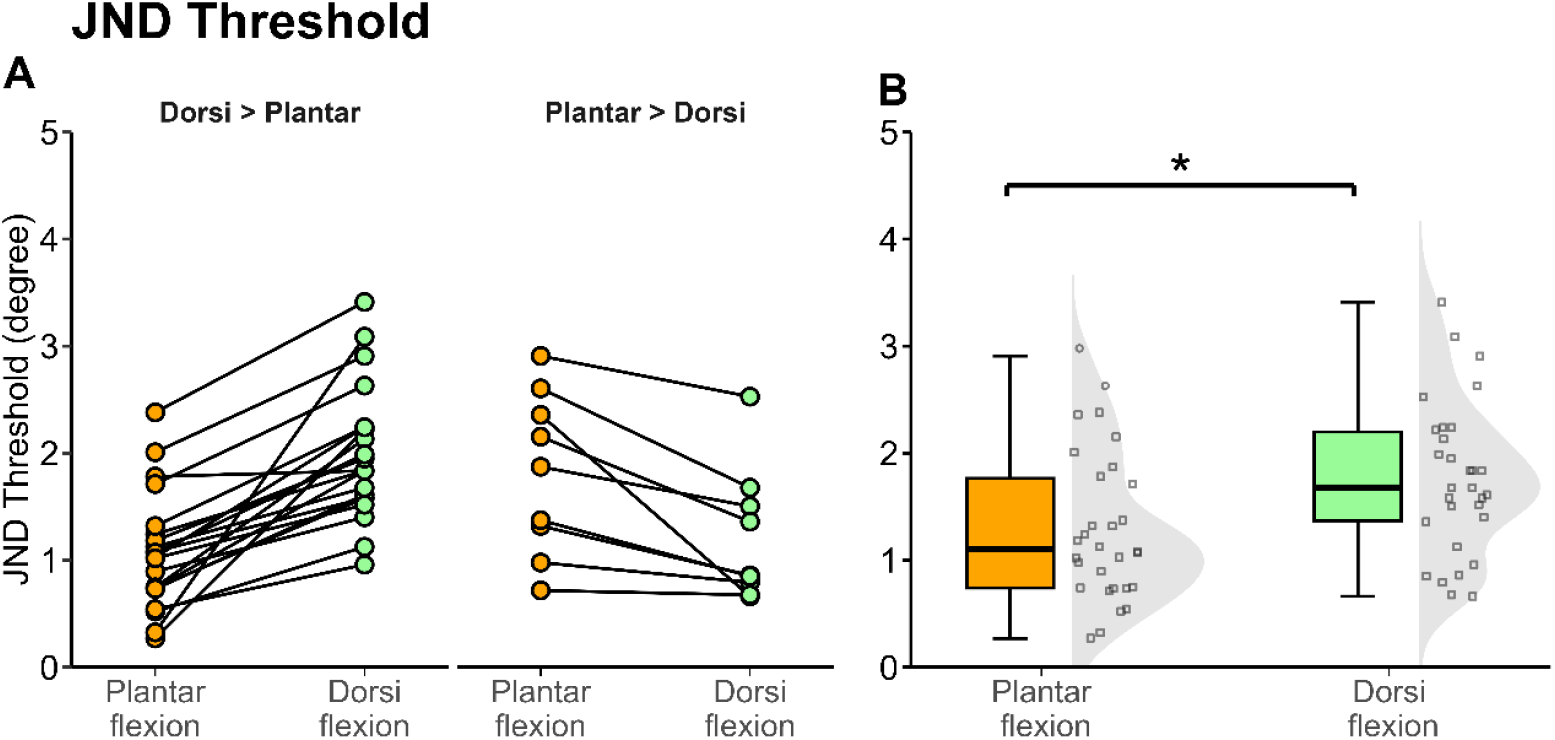
JND Threshold. **(A)** Individual differences between JND thresholds for plantarflexion and dorsiflexion. For the majority of participants (21/30), the JND threshold for plantarflexion was lower than for dorsiflexion. **(B)** The box plot and distribution of JND threshold in plantarflexion and dorsiflexion. The bottom of the box indicates the 25th percentile, the line inside the box marks the median, and the top of the box denotes the 75th percentile. The whiskers extend to 1.5 times the interquartile range of each distribution. Within the half-violin plot, each data point symbolizes an individual threshold value for the ankle. The JND threshold values for plantarflexion were significantly lower than dorsiflexion (* denotes *p* < 0.05).

### Differences in Uncertainty Area between Plantarflexion and Dorsiflexion

To assess differences in proprioceptive precision for plantar-versus dorsiflexed ankle position, we analyzed the individual UA data of all participants. The data in **Figure 3** show that a total of 24/30 (80%) participants exhibited a higher uncertainty in dorsiflexion when compared to plantarflexion. A corresponding Wilcoxon signed-rank test indicated that median UA for plantarflexion was significantly lower when compared to dorsiflexion (*Median* plantarflexion: 0.9° [IQR: 0.9°]; *Median* dorsiflexion: 1.7° [IQR: 1.5°]; *W* = 387, *p* = 0.001). The rank-biserial correlation yielded a moderate to strong effect size (*r*_*b*_= 0.58).

**Figure 3.**
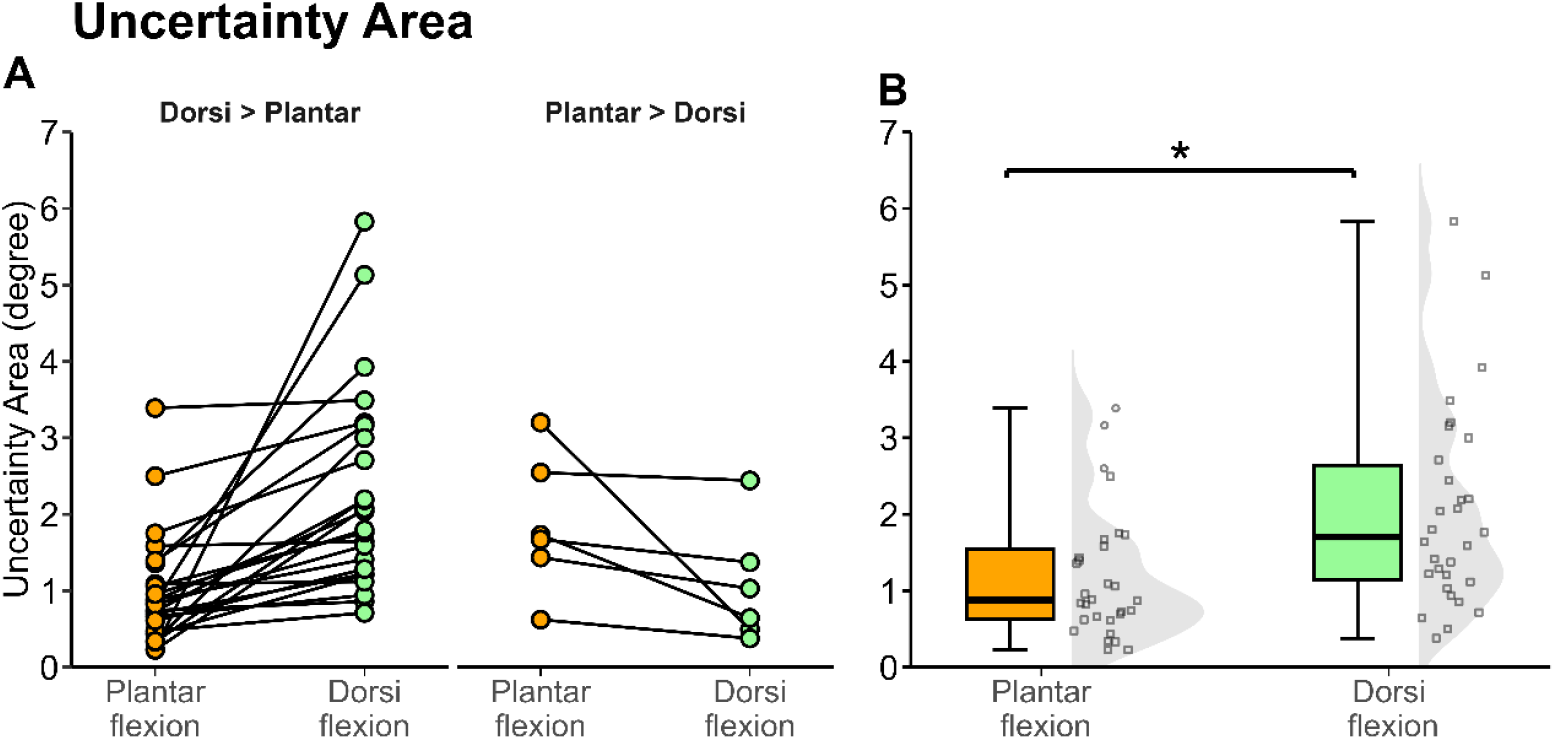
Uncertainty Area. **(A)** Individual differences between UA for plantarflexion and dorsiflexion. For the majority of participants (24/30), the UA for plantarflexion was lower than for dorsiflexion. **(B)** The box plot and distribution of UA threshold in plantarflexion and dorsiflexion. The bottom of the box indicates the 25th percentile, the line inside the box marks the median, and the top of the box denotes the 75th percentile. The whiskers extend to 1.5 times the interquartile range of each distribution. Within the half-violin plot, each data point symbolizes an individual threshold value for the right foot of each participant. The UA values for plantarflexion were significantly lower than dorsiflexion (* denotes *p* < 0.05).

## Discussion

This study examined differences in ankle position sense acuity between plantarflexion and dorsiflexion. We applied an established sensory psychophysics method that yielded two distinct measures to characterize a person’s ability to discriminate ankle positions – a discrimination or JND threshold to capture the magnitude of proprioceptive bias, and the UA to quantify proprioceptive precision, that is, the consistency of sensing a specific joint position over repeated exposure. The main finding of this study is that both measures indicate that ankle position sense acuity for plantarflexion is higher than for dorsiflexion. We discuss possible physiological and biomechanical reasons for this result and how this may affect physical rehabilitation protocols after orthopedic or nervous system injury.

### Difference in Characteristics of Ankle Plantarflexion and Dorsiflexion

A possible reason for the observed difference in proprioceptive acuity for ankle joint plantar-versus dorsiflexion may be the variation in underlying physiological and anatomical structures for both ankle movements. Previous research established that the number and density of muscle spindles, which serve as primary mechanoreceptors for proprioception, are associated with factors such as muscle fiber length and pennation angle (Kissane et al., 2023; Kröger & Watkins, 2021). Ankle plantarflexor muscles, such as soleus and gastrocnemius, are larger and have longer fibers compared to ankle dorsiflexors, such as the tibialis anterior (Brockett & Chapman, 2016; Kissane et al., 2023; Lai et al., 2016). In humans, the mass of the major plantarflexor muscles (e.g., gastrocnemius, soleus) is approximately three to four times larger than for dorsiflexor muscles (extensor digitorum longus, tibialis anterior), but the total number of spindles in major plantarflexors is only about 20% higher than dorsiflexor muscles (Banks, 2006). Thus, structural and size differences between plantar- and dorsiflexor muscles do not provide a conclusive answer to the observed differences in position sense.

In addition to structural factors, the distinct roles of these muscle groups for balance and locomotion may explain the differences in proprioception as the demands on motor control drive the need of the central nervous system for higher resolution proprioceptive feedback. Plantarflexion plays a critical role in activities such as walking, running, and jumping, contributing significantly to balance and stability (Ong et al., 2019). For instance, plantar flexors generate approximately 67% of the total mechanical output of the leg during the propulsion phase of walking and are instrumental in producing forward acceleration during the push-off phase of gait (Montero-Odasso et al., 2005; Ong et al., 2019). These functional demands may necessitate that the proprioceptive signals from plantarflexor muscles have a higher spatial and temporal resolution to meet the requirements for the control of plantarflexion. In contrast, while dorsiflexion is important, it does not require the same level of proprioceptive precision, which the higher UA values found in this study clearly indicate.

Comparisons with other joints, such as the wrist, provide additional insights into proprioceptive mechanisms. Studies on wrist proprioception have shown that flexion/extension movements exhibit higher mechanoreceptor density compared to abduction/adduction movements (Albanese et al., 2021). This aligns with our psychophysical findings on ankle proprioception and underscores the influence of movement direction on proprioceptive acuity. Furthermore, wrists are often involved in fine motor activities, which again underlines that demands on function drive structural differences. This contrast emphasizes the role of functional demands in shaping proprioceptive systems across joints and movement types.

### Limitations

A key limitation of this study is the diversity in the sample. Differences in skeletal muscle composition across ethnic groups, as noted by Silva et al. (2010), suggest that ethnicity may influence proprioception due to variations in muscle spindle characteristics. Additionally, factors such as cultural context, age, gait patterns, and health conditions are relevant to ankle proprioception (Hubbuch et al., 2015; Sertic et al., 2024; Yang et al., 2022).

## Conclusion

This study presents initial evidence about the ankle acuity differences between plantarflexion and dorsiflexion. These differences likely reflect variations in physiological, anatomical, and functional factors. The research highlights the significance of ankle position acuity across different movement directions, providing insights that may enable clinicians to more thoroughly understand ankle joint biomechanics and its role in motor control. This knowledge could contribute to the development of targeted assessments and interventions for proprioceptive deficits, and ultimately for more effective rehabilitation programs for individuals with neurological disorders or orthopedic injuries.

## Data Availability

All data produced in the present study are available upon reasonable request to the authors

## Data Sharing

The data of this study are available upon request by contacting the corresponding author.

## Acknowledgements

This study was supported by internal funds of the University of Minnesota to JK. No Generative AI tools were used for the writing of this manuscript.

